# Data-driven brain atrophy staging in spinocerebellar ataxia type 3

**DOI:** 10.1101/2024.05.29.24307992

**Authors:** Hannah Baumeister, Tamara Schaprian, Philipp Wegner, Monica Ferreira, David Kuegler, Marcondes Cavalcante Franca, Thiago Junqueira Ribeiro de Rezende, Alberto Rolim Muro Martinez, Hong Jiang, Marcus Grobe-Einsler, Berkan Koyak, Demet Önder, Bart van de Warrenburg, Judith van Gaalen, Alexandra Durr, Giulia Coarelli, Matthis Synofzik, Ludger Schöls, Paola Giunti, Hector Garcia-Moreno, Gülin Öz, James M. Joers, Dagmar Timmann, Heike Jacobi, Jeroen de Vries, Peter Barker, Eva-Maria Ratai, Kathrin Reetz, Jon Infante, Jeannette Hübener-Schmid, Thomas Klockgether, ESM MRI study group, DANCER/DELCODE, David Berron, Jennifer Faber

**Author notes:** Correspondence to: Hannah Baumeister, Full address: Leipziger Strasse 44, 39120 Magdeburg, E-Mail, Correspondence may also be addressed to: Jennifer Faber, Full address: Venusberg-Campus 1, 53127 Bonn. D.B. and J.F. contributed equally.

## Abstract

Spinocerebellar ataxia type 3 (SCA3) is the most common autosomal dominant ataxia worldwide. First targeted gene therapy trials have started, offering the intriguing scenario of preventive treatment. SCA3 is associated with progressive regional brain atrophy that starts before clinical manifestation. We aimed to identify the spatiotemporal progression pattern of brain atrophy of SCA3 with a focus on early disease stages.

T1-weighted MRI scans of 300 SCA3 mutation carriers and 317 controls were analyzed. Subtype and Stage Inference (SuStaIn) was used to identify the sequence of volume loss across selected brain regions.

We observed one distinct sequence of brain atrophy events in SCA3 without evidence for the existence of alternative cascades. Atrophy started in the most caudal parts of the brainstem. Almost all preataxic SCA3 mutation carriers clustered in the first atrophy stages. Certainty of sequence estimation was highest for early atrophy stages with prominent involvement of the pons and cerebellar white matter.

Brain atrophy in SCA3 follows a clear and distinct sequence ascending from the lower brainstem with an early involvement of white matter. Knowledge of this sequence might support the stratification of SCA3 mutation carriers with an imminent clinical onset for early interventions.

## Introduction

Spinocerebellar ataxia type 3 (SCA3) is the most common autosomal dominantly inherited ataxia worldwide with a symptom onset in adult life. Clinical hallmarks are a progressive loss of balance and coordination accompanied by slurred speech, leading to disability and premature death. SCA3 is caused by unstable expansions of polyglutamine encoding CAG repeats, resulting in the formation of an abnormally elongated disease protein.^1^ Targeted therapies are being developed, and first safety trials of antisense oligonucleotides (ASOs) have been initiated (https://clinicaltrials.gov, NCT05160558, NCT05822908). Assuming safety and tolerability, such therapies offer the intriguing option of a preventive treatment with the goal of delaying the clinical onset and thus the manifest phase of the disease.

SCA3 is associated with progressive regional brain atrophy mainly affecting the brainstem, cerebellum and basal ganglia, which has been described to develop before clinical manifestation.^2-5^ It has previously been shown that the volume of the pons and cerebellar white matter dropped below the normal range in proximity to the estimated clinical onset, while an increase of the blood-based biomarker neurofilament light (NfL), indicative of ongoing axonal degeneration, was detectable more than a decade before the clinical onset.^6^

The aims of the current study were two-fold. First, we wanted to study whether SCA3 is characterized by a uniform pattern of atrophy development, or whether there are different progression subtypes. Second, we wanted to precisely define the order of regional brain atrophy in the identified pattern(s). For this, we used structural MRI scans from a large data set of 300 SCA3 mutation carriers to train a Subtype and Stage Inference (SuStaIn)^7^ model. This approach enables the identification of distinct atrophy progression patterns from cross-sectional biomarker information using data-driven clustering along with event-based modelling approaches. SuStaIn has been applied previously in samples of memory clinic patients^8^ and various neurodegenerative diseases, including Alzheimer’s disease,^7,9^ epilepsy,^10^ and TAR DNA-binding protein-43 (TDP-43) proteinopathies.^11^

## Methods

### Participants

We included 641 participants, comprising 319 SCA3 mutation carriers and 322 healthy controls (HC), from observational studies at 21 sites worldwide (see Supplementary Table 1). HC were only included for the scaling of MRI-based atrophy markers and were not further analyzed.

### Clinical characteristics of SCA3 mutation carriers

We used the Scale for the Assessment and Rating of Ataxia (SARA)^12^ to assess the presence and severity of ataxia. Manifest ataxia was defined by a SARA sum score of ≥ 3, the term “preataxic” is used for all SCA3 with a SARA sum score < 3.^13^ The SARA sum score was missing for one ataxic SCA3. Neurological symptoms other than ataxia were assessed using the Inventory of Non-Ataxia Signs (INAS).^14^ INAS count was available for 188, and individual INAS items for 113 cases. Clinical disease stages were delineated based on self-reported gait status as follows: stage 0 (“normal”, no gait dif?culties), stage 1 (“gait difficulties”, reported presence of gait dif?culties), stage 2 (“walking aid”, loss of independent gait, as de?ned by permanent use of a walking aid or reliance on a supporting arm), stage 3 (“wheelchair”, reliance on wheelchair, as de?ned by permanent use of a wheelchair).^15^ Data on clinical disease stages were available for 130 participants.

Repeat lengths of the expanded and normal alleles were determined at the Institute for Medical Genetics and Applied Genomics of the University of Tübingen (*n*=91), Germany, or taken from medical records (*n*=197). No information about repeat lengths of the expanded allele was available for twelve SCA3. Age of onset was defined as the reported first occurrence of gait disturbances and estimated for SCA3 without this information (see Supplementary methods).

### Image analysis

Available T1-weighted MRI scans were visually inspected and screened for motion artifacts or incomplete coverage. The majority of included scans (*n* = 501, 82.4%) were acquired with a 1mm isotropic resolution. The remaining scans were resampled to a 1mm isotropic resolution. Images were N4 bias field corrected (Advanced Normalization Tools, v2.1).^16^ FreeSurfer v6.0 was used to obtain volumes of the cerebrum and brainstem as well as the estimated total intracranial volume.^17,18^ Cerebellar sub-segmentation was performed using CerebNet.^3^ For all hemispheric volumes, left and right hemispheric volumes were summed.

In line with our goal of developing a disease progression model that is particularly sensitive to possible volumetric changes before ataxia onset, we focused on volumes that differed significantly between preataxic SCA3 and HC either in the literature^2-4,19^ or the present cohort (Supplementary methods). We applied a regression-based approach to correct all regional volumes for their relationship with estimated total intracranial volume in the healthy control group.^20^

### Subtype and Stage Inference

SuStaIn was implemented in python v3.9. A detailed methodological description of SuStaIn was given by Young et al.^7^ In the present study, only SCA3 mutation carriers were included in the SuStaIn analysis. All regional volumes were *z*-standardized to the HC group and corrected for their association with age and sex in HCs. Hence, *z*-scores can be interpreted as the age-and sex-adjusted standard deviation (*SD*) differences from the HC group.

We chose *z*∈{−1, −2, −3} as thresholds for atrophy events representing increasing severity. As SuStaIn assumes biomarkers to monotonously increase with disease progression, we inverted *z*-scores for modelling but report original values where lower values represent declining volumes for more intuitive understanding. We trained SuStaIn models to estimate *c*∈{1, 2, 3, 4, 5} atrophy progression sequences of 27 atrophy events, derived from the combination of three *z*-thresholds and nine regional volumes. We refer to the positions of these events within the event sequence as SuStaIn stages, an ordinally scaled measure of atrophy progression. While continuous modelling occurred across SuStaIn stages, their scaling does not allow for an interpretation as continuously scaled time-equivalents. In other words, the time interval between two stages is not equidistant.

### Statistical analysis

Statistical analyses were performed in R v4.2.2. The pseudo-longitudinal change of regional volumes across SuStaIn stages was modelled using monotone penalized cubic regression splines. A Welch Two Sample *t*-test was used to compare mean SuStaIn stages of preataxic and ataxic participants and of participants with an without individual non-ataxia signs. Bhattacharyya coefficients were calculated to quantify the overlap of atrophy progression sequences across atrophy events and cross-validation folds. They can range from 0 to 1 with higher values reflecting an increasing overlap of atrophy sequences across folds. We performed analyses of variance with Tukey’s Honest Significant Difference *post-hoc* comparisons to test for differences in mean SuStaIn stage across clinical disease stages. Spearman rank correlations were used to test if SuStaIn stage was correlated with time from ataxia onset, SARA sum scores, and INAS count. *P*-values were corrected for false discovery rate (FDR) where appropriate.

## Results

Twenty-four participants had to be excluded due to scanning artifacts or incomplete clinical information. Demographic and clinical data of the remaining 300 SCA3 mutation carriers and 317 healthy controls are summarized in Table 1.

**Table 1.**
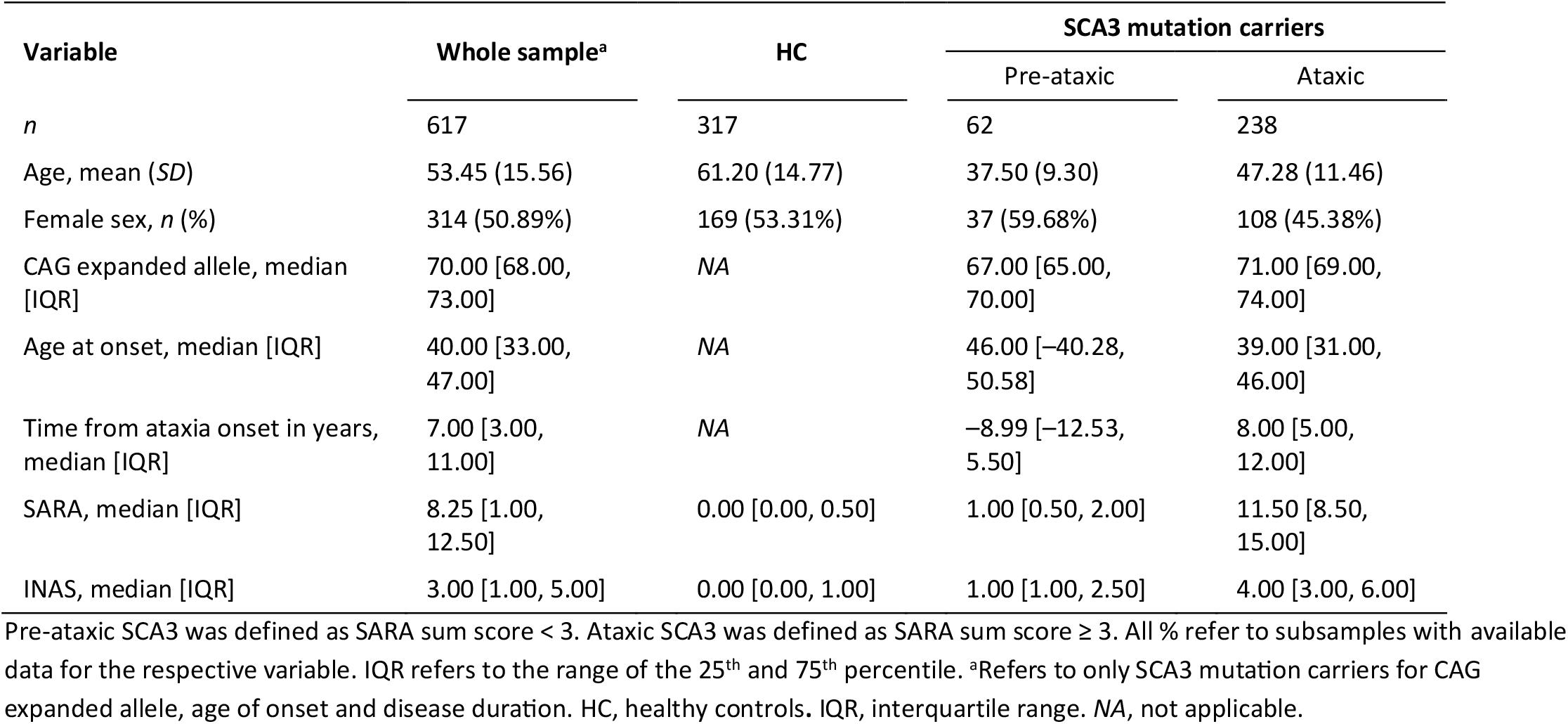
Characteristics of the analyzed sample.

### Regional atrophy in SCA3 progresses along one unambiguous spatiotemporal cascade

The following volumes of the cerebrum, cerebellum, and brainstem were included in the SuStaIn model: medulla oblongata, pons, midbrain, cerebellar white matter, anterior, inferior posterior, and flocculonodular lobe of the cerebellum as well as pallidum and thalamus. Ten-fold cross-validation of SuStaIn models of up to five distinct atrophy sequences revealed one unambiguous event sequence of reduced volumes at different degrees among SCA3 mutation carriers (Figure 1A,B). This is strong evidence against the existence of multiple cascades of brain atrophy in SCA3 mutation carriers. High Bhattacharyya coefficients across atrophy events and cross-validation folds indicated an excellent internal robustness of the proposed event sequence (mean = 0.92; *SD* = 0.10; Figure 2A). Subject-level classifications were made with highest certainty in early SuStaIn stages where the majority of preataxic participants were clustered (Figure 2B, C). Further supporting results of cross-validation and model selection are provided in the Supplementary Results.

**Figure 1.**
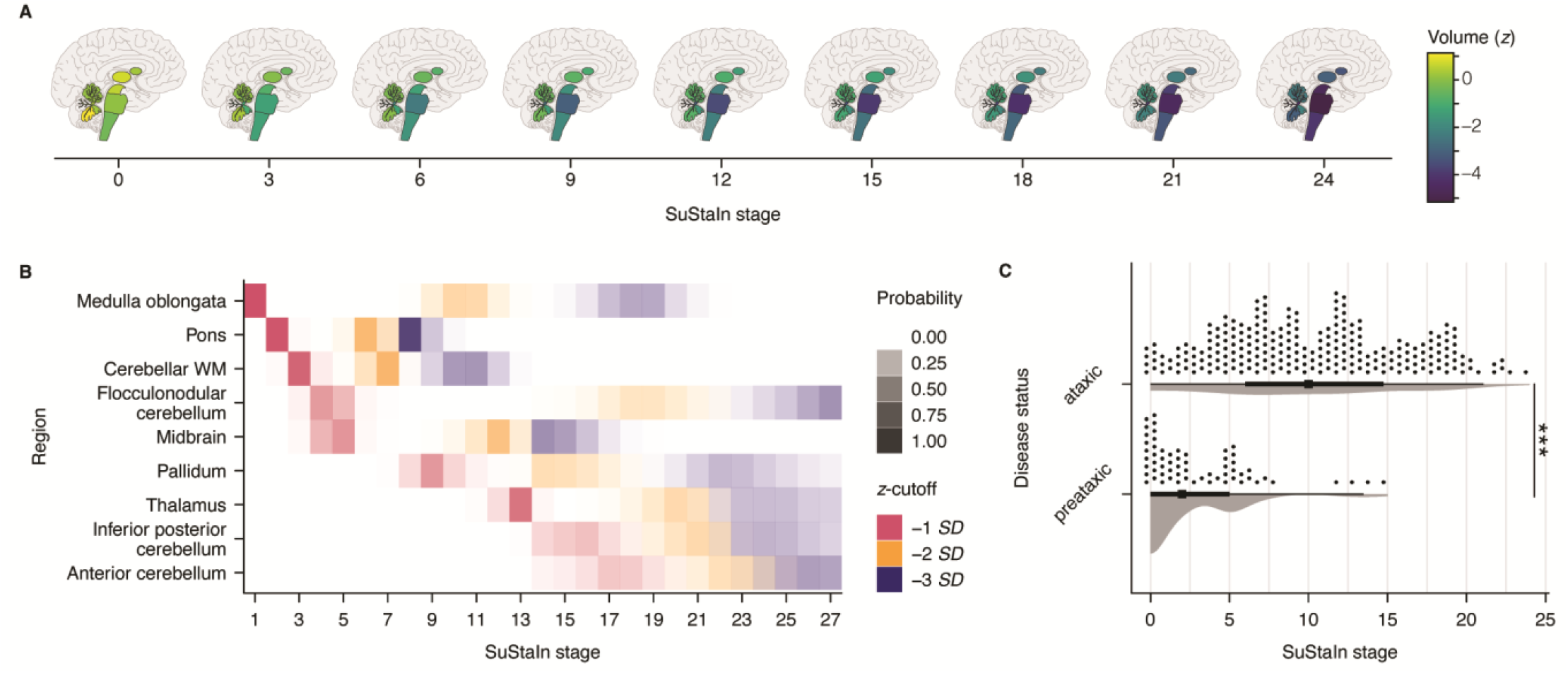
Sequence of brain atrophy in SCA3 mutation carriers. SuStaIn was used to estimate the progression sequence of atrophy in SCA3 mutation carriers. (**A**) Pictograms dipict the obtained sequence. The displayed *z*-scores were predicted from monotone penalized cubic regression splines modelling the relationship of *z*-scores of each volume and SuStaIn stage (see also Supplementary Figure 2). (**B**) The positional variance diagram depicts the certainty with which each atrophy event was located within the entire atrophy sequence. Increasing opaqueness represents a higher certainty of an atrophy event occurring in the displayed location within the sequence. (**C**) Distributions of SuStaIn stages in the present sample stratified by disease status. \*\*\**P* < .001.

**Figure 2.**
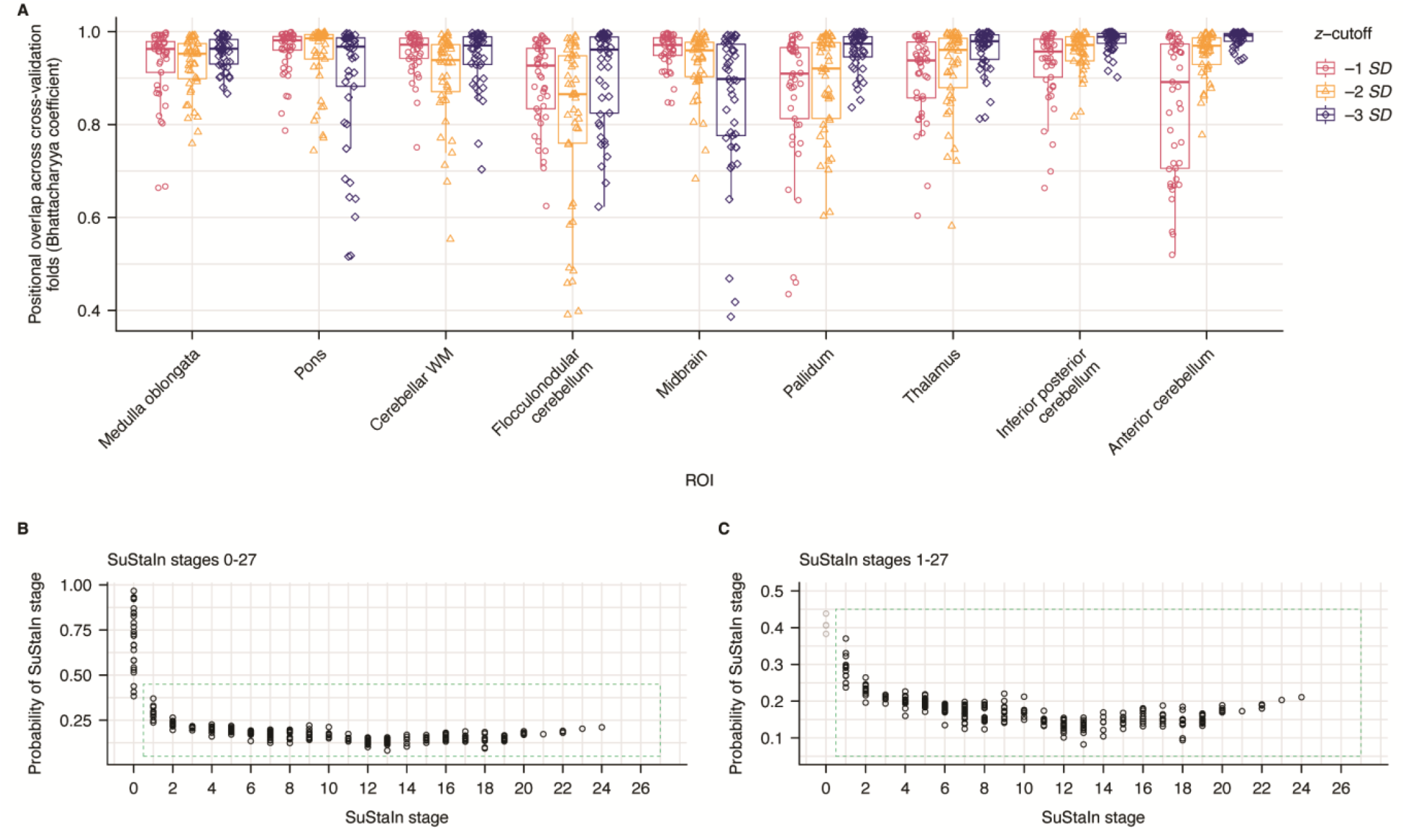
Model performance in ten-fold cross-validation and subject-level classification certainties. (**A**) Bhattacharyya coefficients were calculated for each atrophy event across pairs of cross-validation folds. The overall high values indicate high model robustness and generalizability within the sample. (**B**, **C**) On a participant level, probabilities of the assigned SuStaIn stage were highest in early SuStaIn stages.

The identified sequence of brain atrophy is visualized in Figure 1A,B (see also Supplementary Figure 2). In brief, initial volume loss (–1 *SD*) occurred in the medulla oblongata, pons, cerebellar white matter, the flocculonodular cerebellum, and the midbrain. Notably, further atrophy (–2 and –3 *SD*) most rapidly progressed in the cerebellar white matter and the pons (Figure 1B). The mean SuStaIn stage was significantly higher in ataxic (10.26 ± 5.76) than in preataxic SCA3 (3.15 ± 3.59; *t*(152.47) = –12.07, *P* < .001; Figure 1C). 55 out of 62 preataxic SCA3 mutation carriers (91.7%) were grouped within the first seven SuStaIn stages.

### SuStaIn stages correlate with disease duration and ataxia severity

Mean SuStaIn stage differed among the different clinical disease stages (*F*(3, 126) = 21.47, *η*^2^ = 0.34, *P* < .001). Participants in stages 1 (“gait difficulties”) and 2 (“walking aid”) were each classified with higher SuStaIn stages than those in previous stages. Wheelchair-reliant patients were classified with higher SuStaIn stages than those in stage 0 (“normal”; Figure 3A, Supplementary Table 4).

**Figure 3.**
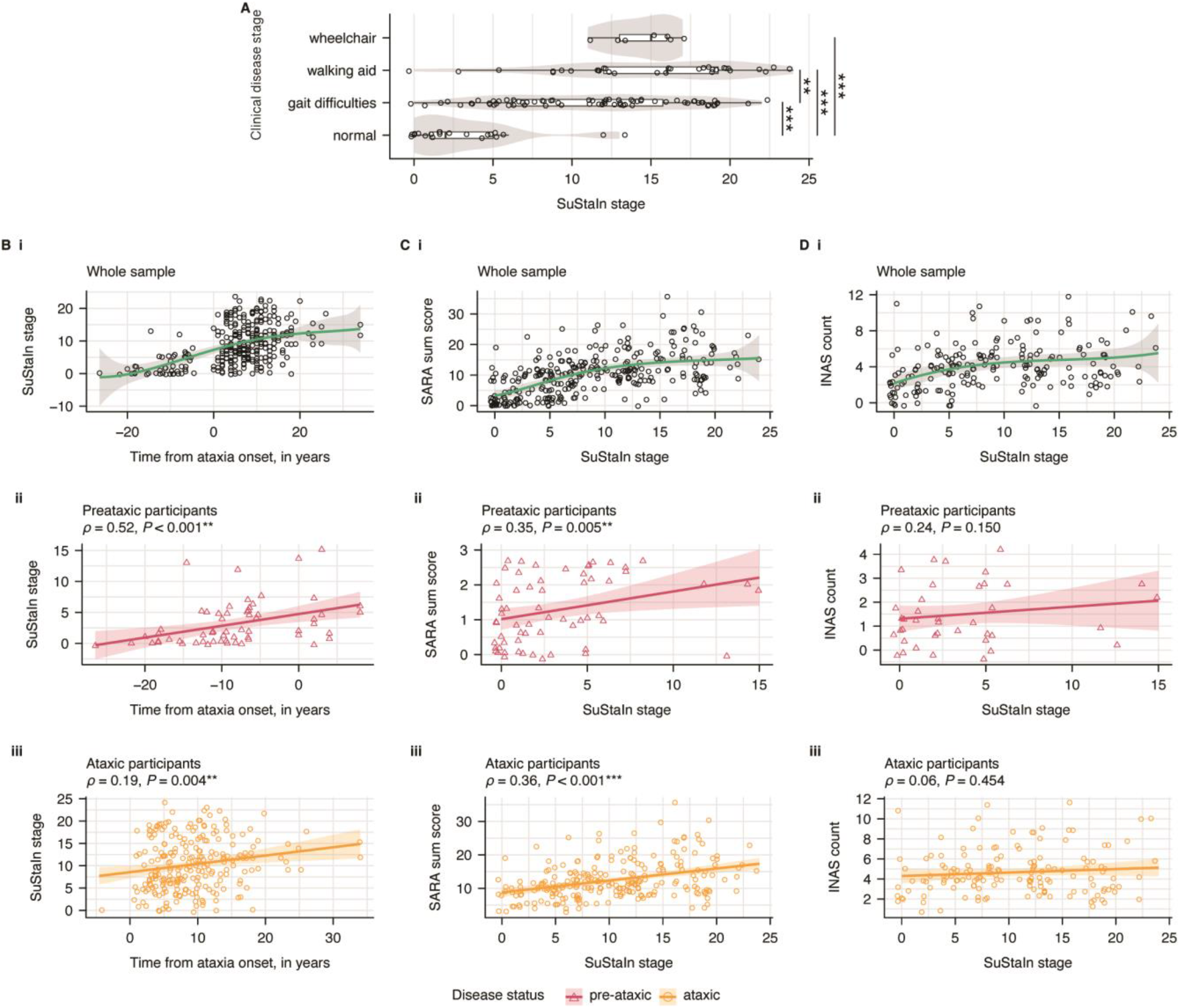
Relationships of SuStaIn stage with time from ataxia onset, ataxia severity, and INAS count. (**A**) Mean SuStaIn stages of SCA3 mutation carriers across increasing degrees of gait deterioration were significantly higher than without gait disturbance. (**B**) The relationship of SuStaIn stages and time from ataxia onset was estimated using (**B i**) natural cubic regression splines for the whole sample of SCA3 mutation carriers and linear correlations for (**B ii**) preataxic and (**B iii**) ataxic SCA3 separately. (**C**,**D**) The same analysis steps were used to explore the relationships of SARA sum scores and INAS count with SuStaIn stage. Note that, in all plots, SuStaIn stage values were jittered on the respective axes. *P*-values in (**A**) are FDR-corrected. \**P* < .05. \*\**P* < .01. \*\*\**P* < .001.

Within the entire sample, SuStaIn stage continuously increased with disease duration (Figure 3Bi). However, when considering preataxic and ataxic SCA3 separately, the correlation of participants’ disease duration with SuStaIn stage was strong in preataxic participants (*ρ* = 0.52, *P* < 0.001) and weak in ataxic participants (*ρ* = 0.19, *P* = 0.004; Figures 3Bii,iii).

The association of SARA scores and SuStaIn stage followed an asymptotic curve with SARA scores rapidly increasing in early SuStaIn stages and approaching a plateau in the second half of the atrophy cascade (Figure 3Ci). Linear correlations of SARA scores and SuStaIn stage were positive and significant in both preataxic (*ρ* = 0.35, *P* = 0.005) and in ataxic (*ρ* = 0.36, *P* < 0.001) SCA3 (Figures 3Cii,iii).

We found no correlation of INAS count and SuStaIn stage in both preataxic (*ρ* = 0.24, *P* = 0.150) and ataxic (*ρ* = 0.06, *P* = 0.454) SCA3 (Figure 3Di–iii). However, when investigating non-ataxia signs individually, SuStaIn stages were significantly elevated in participants with spasticity, fasciculations, brainstem oculomotor signs, urinary dysfunction, resting tremor, and hypereflexia (test statistics reported in Supplementary Table 5; see also Supplementary Figure 3).

## Discussion

We used SuStaIn modelling to study the spatiotemporal order of brain atrophy in SCA3. Our data set of 300 T1-weighted MRI scans of SCA3 mutation carriers covered a broad range of the disease course including preataxic mutation carriers with an estimated time of up to more than 25 years before ataxia onset to patients with a reported disease duration of more than 30 years. With this data-driven approach, we – for the first time – demonstrated a distinct sequence of atrophy events without evidence of multiple atrophy subtypes. In this sequence, atrophy ascends from the lower brainstem with an early and strong involvement of the cerebellar white matter. Pons and cerebellar white matter showed the fastest volume decrease. Over 90% of preataxic SCA3 were grouped within the first seven out of 27 SuStaIn stages.

Our study supports that SCA3 is characterized by a uniform pattern of atrophy development. This distinguishes SCA3 from other neurodegenerative diseases, such as Alzheimer’s disease and TDP-43 proteinopathies.^11,21^ This clear difference may be explained by the single genetic cause of SCA3 and younger mean age but is not self-evident, as various clinical subtypes of SCA3 have been described.^22-25^ Moreover, many of the neurodegenerative diseases with evident biological heterogeneity occur in older age. It is possible that this heterogeneity is partially driven by age-related co-pathologies, which are less common in the relatively young patients with SCA3.

While previous studies showed an early involvement of brainstem and cerebellar white matter atrophy in SCA3,^2-6,19,26,27^ we here provide evidence of brain atrophy ascending from the lower brainstem via the pons and cerebellar white matter to the cerebellar cortex, midbrain, thalamus, and basal ganglia. This sequence, with a start in the medulla oblongata, may suggest that the actual origin of pathology of SCA3 lies in the spinal cord. Unfortunately, the available data did not allow a reliable assessment of the spinal cord. Accordingly, there is a need for MRI studies including the spinal cord in SCA3.

Notably, the identified sequence follows anatomical connections, which raises the question of the underlying pathological mechanisms. In principle, this observation is compatible with transneuronal spreading of an abnormal disease protein, as put forward in other neurodegenerative diseases.^28^ To date, there is no clear evidence of a spreading pathology in SCAs.^29^ Meanwhile, these anatomical connections are formed by white matter tracts and there is increasing evidence for a crucial role of oligodendroglia in SCA3.^30,31^

Overall clinical deterioration, indicated by clinical disease stages, was associated with higher SuStaIn stages, except for the latest clinical disease stage (wheelchair-reliant patients). This group only comprises six participants and most likely represents a selection of less affected patients who are still able to participate in study visits. In ataxic SCA3, SuStaIn stage was more strongly correlated with ataxia severity, as measured by SARA, than with disease duration. This result suggests that ataxia severity is determined by the extent of brain structural changes. A similar correlation has been previously shown for brainstem volume in SCA3.^32^ In preataxic mutation carriers, in whom SARA scores vary only in a small (normal) range, SuStaIn stages were more strongly correlated with time from ataxia onset than with SARA scores. The INAS count was not correlated with SuStaIn stages, suggesting that accompanying non-ataxia signs in general are not strongly determined by the volumes of the studied brain regions. This is not surprising, as INAS is composed of a variety of signs, including those that are due to peripheral neuropathy, such as areflexia. Meanwhile, certain signs that clearly have a central origin, such as spasticity and brainstem oculomotor signs, were associated with higher SuStaIn stages.

The proposed model most confidently classified SCA3 mutation carriers in early SuStaIn stages. While we cannot be certain about the exact reasons for this result, there are various plausible explanations. First, even though our sample was large for SCA3 being a rare disease, we had to limit the number of considered regions to reduce model complexity. We focused on those regions in which atrophy could already be detected in the preataxic disease stage. Second, higher model certainty in early SuStaIn stages may be a result of most participants clustering in these stages and, thus, increasing statistical power. Third, brain atrophy may progress in a strictly uniform fashion in early disease stages but may then individualize.

Overall, the *a priori* assumptions and constrains to our model were selected to increase model sensitivity in early, preataxic disease stages of SCA3, as this is of particular interest for future therapeutic developments. Given the results, including the high model certainty in early atrophy stages and the high correlation with time from ataxia onset in preataxic mutation carriers, this strategy was successful. Meanwhile, conclusions regarding the late symptomatic phase should be drawn with caution.

The proposed model demonstrated high robustness in our internal cross-validation procedure. Future studies using external validation data are needed to determine whether this model is generalizable to novel test data that were not used for training.^33^ Aside external validation, longitudinal studies are needed to assess if atrophy, indeed, progresses along the proposed pseudo-longitudinal sequence.

In conclusion, we provide evidence that atrophy in SCA3 follows a clear and distinct sequence ascending from the lower brainstem with an early affection of white matter. The vast majority of preataxic SCA3 mutation carriers were classified in a portion of this sequence that was characterized by atrophy progressing rapidly in the pons and the cerebellar white matter. Consequently, these volumes may be useful stratification markers for the imminent ataxia onset in preataxic SCA3 with relevance for the design of future preventive trials.

## Data Availability

All data produced in the present study are available upon reasonable request to the authors.

## Acknowledgements

We would like to acknowledge the institutional and personal contributions and support of this work by the Imaging Platform ICM (CENIR) and Hortense Hurmic, research assistant at the ICM, Paris; Khalaf O. Bushara from the Department of Neurology, University of Minnesota, Minneapolis, MN, USA, Diane Hutter, Center for Magnetic Resonance Research, Department of Radiology, University of Minnesota, Minneapolis, MN, USA, Andreas Thieme, Friedrich Erdlenbruch, Thomas M. Ernst from the Department of Neurology and Center for Translational Neuro- and Behavioral Sciences, University Hospital Essen, University of Duisburg-Essen, Duisburg, Germany as well as Jeremy Schmahmann and Jason Macmore from the Ataxia Center, Laboratory for Neuroanatomy and Cerebellar Neurobiology, Massachusetts General Hospital and Harvard Medical School, Boston, MA, USA. This publication is an outcome of ESMI, an EU Joint Programme - Neurodegenerative Disease Research (JPND) project (see www.jpnd.eu). The project is supported through the following funding organisations under the aegis of JPND: Germany, Federal Ministry of Education and Research (BMBF; funding codes 01ED1602A/B); Netherlands, The Netherlands Organisation for Health Research and Development; United Kingdom, Medical Res earch Council (MR/N028767/1). At the US sites this work was in part supported by the National Ataxia Foundation and the National Institute of Neurological Disorders and Stroke (NINDS) grant R01NS080816. The Center for Magnetic Resonance Research is supported by the National Institute of Biomedical Imaging and Bioengineering (NIBIB) grant P41 EB027061, the Institutional Center Cores for Advanced Neuroimaging award P30 NS076408 and S10 OD017974 grant. JF received consultancy frees from Vico Therapeutics, unrelated to the present manuscript, and funding from the Advanced Clinician Scientist Programme (ACCENT, funding code 01EO2107, the ACCENT Program is funded by the German Federal Ministry of Education and Research (BMBF)) as well as a fellow of the Hertie Network of Excellence in Clinical Neuroscience. TK is receiving research support from the Bundesministerium für Bildung und Forschung (BMBF) and Servier. MCF is funded by Friedreich’s research alliance (FARA, USA) and Fundação de Amparo à pesquisa do Estado de São Paulo (grants # 2021/06739-4 and 2013/07559-3). TJRR is funded by Friedreich’s research alliance (FARA, USA). No further conflicting interests were disclosed related to this manuscript.

## Supplementary data

### Supplementary methods

#### Estimation of the age of onset

Age of onset was defined as the reported first occurrence of gait disturbances and was missing for 15 SCA3 already experiencing gait disturbances. Here, we estimated the age of onset based on the CAG repeat length.^34^ For SCA3 mutation carriers without gait difficulties (i.e., at clinical stage 0), the age of onset was estimated based on CAG repeat length and the current age resulting in negative values giving the estimated time to ataxia onset.^34^ This was not possible for one participant due to missing genetic information.

##### A priori selection of volumes as SuStaIn input

Volumes were selected based on empirical evidence from the present sample and a literature review. In the empirical selection process, various volumes taken from the present dataset were compared across healthy controls, as well as preataxic and ataxic SCA3 mutation carriers. A full list of volumes is given in Supplementary Table 2. For all hemispheric volumes, the volumes of the left and right hemispheres were summed. For the group statistics of the present cohort, all volumes were divided by a participant’s eTIV, and subsequent group statistics were based on these relative values. We conducted an analysis of variance (ANOVA) to assess statistical differences of mean volumes between the groups of preataxic and ataxic SCA3 mutation carriers as well as healthy controls. For *post-hoc* testing we applied Tukey’s Honest Significant Difference testing and corrected *p*-values for family-wise error rate.

In addition to this empirical approach, we also conducted a literature review. Since structural changes in preataxic SCA3 mutation carriers are mainly influenced by the time from ataxia onset with an expected increase in changes with increasing proximity to clinical onset,^4,6^ we also included volumes that appear relevant for the present study based on the existing literature.

##### SuStaIn modelling and model selection

We used internal ten-fold cross-validation to determine the optimal number of subtypes given the training data. To determine the optimal number of subtypes, we used the cross-validation information criterion (CVIC) and mean out-of-sample log-likelihood from each model establishing *c*∈{1,2,3,4,5} atrophy progression sequences. A more complex model (a model with *c*-subtypes) was preferred over a simpler model (a model with *c* − 1 subtypes) only if (a) CVIC_*c*_ − CVIC_*c*−1_ < −3, and (b) log-likelihood_*c*_ − log-likelihood_*c*−1_ > 6.

### Supplementary results

#### Volume selection

The results of diagnostic group comparisons across regional volumes are shown in Supplementary Table 2. In combination with the the literature review, we decided to include nine volumes as summarized in Supplementary Table 3.

#### SuStaIn model selection

Given the pre-defined criteria for model selection, the model assuming *c* = 1 subtypes of atrophy progression most efficiently explained the training data. The CVIC and out-of-sample log-likelihood statistics obtained from ten-fold cross-validation are plotted in Supplementary Figure 1.

## Supplementary tables

**Supplementary Table 1.** Overview of scanners and sequences. *NA* = missing data. T, Tesla. TR, repetition time. TE, echo time. TI, inversion time

| Country | Site | Number of scans | Scanner vendor | Field strength, T | TR, ms | TE, ms | TI, ms | Resolution, mm |  |  |
| --- | --- | --- | --- | --- | --- | --- | --- | --- | --- | --- |
|  |  |  |  |  |  |  |  | x | y | z |
| Brazil | Campinas | 53 | Phillips, Achieva | 3 | 7.00 | 3201 | NA | 1.00 | 1.00 | 1.00 |
| China | Changsha | 50 | SIEMENS, Prisma | 3 | 2300 | 2.98 | 900 | 0.90 | 0.94 | 0.94 |
| France | Paris | 2 | GE, SIGNA HDX | 3 | 5.96 | 2.532 | 380 | 0.89 | 0.89 | 1.00 |
|  |  | 1 | GE, Optima MR360 | 1.5 | 7.94 | 4.20 | NA | 0.50 | 0.50 | 1.40 |
|  |  | 1 | GE, SIGNA HDX | 3 | 6.61 | 2.828 | NA | 1.00 | 1.00 | 1.20 |
|  |  | 1 | GE, Optima MR450w | 1.5 | 9.28 | 4.20 | NA | 0.50 | 0.50 | 1.20 |
|  |  | 19 | SIEMENS, TRIO | 3 | 2300 | 2.98 | 900 | 1.00 | 1.00 | 1.10 |
| Germany | Aachen | 6 | SIEMENS, Prisma | 3 | 2500 | 4.37 | 1100 | 1.00 | 1.00 | 1.00 |
|  | Berlin | 54 | SIEMENS, TimTrio | 3 | 2500 | 4.33 | 1100 | 1.00 | 1.00 | 1.00 |
|  |  | 60 | SIEMENS, Prisma fit | 3 | 2500 | 4.37 | 1100 | 1.00 | 1.00 | 1.00 |
|  | Bonn | 112 | SIEMENS, Skyra | 3 | 2500 | 4.37 | 1100 | 1.00 | 1.00 | 1.00 |
|  |  | 2 | Philips, Intera | 1.5 | 15.31 | 3.6 | NA | 1.00 | 1.00 | 1.00 |
|  |  | 8 | Philips, Intera | 3 | 8.29 | 3.8 | NA | 1.00 | 1.00 | 1.00 |
|  | Essen | 12 | SIEMENS, Biograph | 3 | 2500 | 4.37 | 1100 | 1.00 | 1.00 | 1.00 |
|  | Frankfurt | 9 | SIEMENS, Verio | 3 | 1500 | 2.93 | 900 | 0.50 | 0.50 | 1.00 |
|  | Goettingen | 4 | SIEMENS, TrioTim | 3 | 2500 | 4.33 | 1100 | 1.00 | 1.00 | 1.00 |
|  | Heidelberg | 11 | SIEMENS, TRIO | 3 | 2501 | 4.33 | 1100 | 1.00 | 1.00 | 1.00 |
|  | Magdeburg | 24 | SIEMENS, Verio | 3 | 2500 | 4.37 | 1100 | 1.00 | 1.00 | 1.00 |
|  | Munich | 21 | SIEMENS, Skyra | 3 | 2500 | 4.37 | 1100 | 1.00 | 1.00 | 1.00 |
|  | Rostock | 27 | SIEMENS, Skyra fit | 3 | 2500 | 4.37 | 1100 | 1.00 | 1.00 | 1.00 |
|  | Tuebingen | 8 | SIEMENS, Prisma | 3 | 2500 | 4.37 | 1100 | 1.00 | 1.00 | 1.00 |
|  |  | 1 | SIEMENS, Sonata | 1.5 | 20.00 | 5 | NA | 0.90 | 0.90 | 0.90 |
|  |  | 11 | SIEMENS, Avanto | 1.5 | 1900 | 3.93 | 1100 | 1.00 | 1.00 | 1.00 |
|  |  | 1 | SIEMENS, Skyra | 3 | 2300 | 23.2 | 900 | 0.90 | 0.90 | 0.90 |
|  |  | 1 | SIEMENS, Sonata | 1.5 | 1150 | 32.2 | NA | 0.48 | 0.48 | 1.00 |
|  |  | 1 | SIEMENS, Sonata | 1.5 | 1430 | 36.8 | NA | 0.50 | 0.50 | 1.00 |
|  |  | 1 | SIEMENS, Avanto | 1.5 | 1900 | 30.9 | 1100 | 1.00 | 1.00 | 1.00 |
|  |  | 13 | SIEMENS, Skyra | 3 | 2300 | 11720 | 900 | 0.90 | 0.90 | 0.90 |
| Spain | Santander | 2 | Philips, Achieva | 3 | 15.00 | 5 | NA | 0.90 | 0.90 | 1.00 |
| The Netherlands | Groningen | 7 | SIEMENS, Prisma | 3 | 2500 | 4.37 | 1100 | 1.00 | 1.00 | 1.00 |
|  | Nijmegen | 19 | SIEMENS, Skyra | 3 | 2500 | 4.37 | 1100 | 1.00 | 1.00 | 1.00 |
|  |  | 1 | SIEMENS, Avanto | 1.5 | 1900 | 2.55 | 1100 | 1.00 | 1.00 | 1.00 |
|  |  | 1 | SIEMENS, Avanto | 1.5 | 1900 | 2.55 | 1100 | 0.90 | 0.90 | 1.00 |
|  |  | 24 | SIEMENS, Trio | 3 | 2300 | 3.03 | 1100 | 1.00 | 1.00 | 1.00 |
| UK | London | 21 | SIEMENS, Prisma | 3 | 2500 | 4.37 | 1100 | 1.00 | 1.00 | 1.00 |
| US | Baltimore | 7 | SIEMENS, Prisma fit | 3 | 2500 | 4.37 | 1100 | 1.00 | 1.00 | 1.00 |
|  | Boston | 3 | SIEMENS, TrioTim | 3 | 2500 | 4.33 | 1100 | 1.00 | 1.00 | 1.00 |
|  | Minnesota | 18 | SIEMENS, Prisma Fit | 3 | 2500 | 4.37 | 1100 | 1.00 | 1.00 | 1.00 |

**Supplementary Table 2.** Test statistics of Tukey *post-hoc* comparisons of mean volumes for across different patient groups.

| Volume | Preataxic SCA3<br>< HC | Ataxic SCA3<br>< preataxic SCA3 | Ataxic SCA3<br>< HC |
| --- | --- | --- | --- |
| **Brainstem** |  |  |  |
| Medulla oblongata | *** | *** | *** |
| Pons | *** | *** | *** |
| Midbrain | * | *** | *** |
| **Cerebellum** |  |  |  |
| Cerebellar WM | * | *** | *** |
| Anterior cerebellum |  |  | * |
| Superior posterior cerebellum |  | *** | *** |
| Inferior posterior cerebellum | * | *** |  |
| Flocculonodular cerebellum |  | *** | *** |
| Vermis |  | ** |  |
| Cerebellar GM |  | *** | *** |
| **Basal ganglia and deep gray matter** |  |  |  |
| Thalamus | * | *** | *** |
| Caudate |  | * | ** |
| Pallidum |  | *** | *** |

\* $P_{FWE} < .05$ . \*\* $P_{FWE} < .01$ . \*\*\* $P_{FWE} < .001$ . WM, white matter. GM, gray matter.

**Supplementary Table 3.** Overview of rationales for incorporating individual volumetric markers in SuStaIn model training.

| ROI | Empirical evidence<sup>a</sup> | Chandrasekaran *et al.* (2023)<sup>2</sup> | Faber *et al.* (2021)<sup>4</sup> | Faber *et al.* (2022)<sup>3</sup> |
| --- | --- | --- | --- | --- |
| Medulla oblongata | X | X | X |  |
| Pons | X |  | X |  |
| Midbrain | X |  |  |  |
| Cerebellar WM | X |  |  | X |
| Anterior cerebellum | X |  | X |  |
| Inferior posterior cerebellum | X |  |  |  |
| Flocculonodular cerebellum |  | X |  | X |
| Pallidum |  |  | X |  |
| Thalamus | X |  |  |  |

<sup>a</sup>In the present cohort and partly overlapping with the literature. WM, white matter.

**Supplementary Table 4.** Test statistics of Tukey *post-hoc* comparisons of mean SuStaIn stages for participants at different levels of gait disturbances.

| Contrast | *b* | *SE* | *t* | *P*<sub>FDR</sub> |
| --- | --- | --- | --- | --- |
| gait disturbance *versus* normal | 7.93 | 1.32 | 6.00 | < .001*** |
| walking aid *versus* normal | 11.59 | 1.49 | 7.77 | < .001*** |
| wheelchair *versus* normal | 11.14 | 2.32 | 4.81 | < .001*** |
| walking aid *versus* gait disturbance | 3.66 | 1.13 | 3.23 | .002** |
| wheelchair *versus* gait disturbance | 3.21 | 2.10 | 1.53 | .155 |
| wheelchair *versus* walking aid | −0.45 | 2.22 | −0.20 | .841 |

\* $P_{FDR} < .05$ . \*\* $P_{FDR} < .01$ . \*\*\* $P_{FDR} < .001$ .

**Supplementary Table 5.** Test statistics from Welch two-sample *t*-tests comparing mean SuStaIn stage in SCA3 mutation carriers stratified by presence of different non-ataxia signs.

| INAS item | *n*<sub>+</sub> (*n*<sub>total</sub>) | Mean |  | SD |  | df | *t* | *P* | *P*<sub>FDR</sub> |
| --- | --- | --- | --- | --- | --- | --- | --- | --- | --- |
|  |  | – | + | – | + |  |  |  |  |
| Areflexia | 53 (112) | 10.61 | 11.85 | 7.38 | 5.56 | 106.82 | –1.01 | .315 | .388 |
| Hyperreflexia | 37 (113) | 10.09 | 13.22 | 6.49 | 6.44 | 71.90 | –2.41 | .018* | .049* |
| Extensor plantar response | 8 (111) | 11.10 | 12.50 | 6.72 | 5.15 | 8.96 | –0.72 | .488 | .520 |
| Spasticity | 30 (113) | 9.67 | 15.10 | 6.54 | 5.04 | 66.36 | –4.65 | < .001*** | < .001*** |
| Paresis | 20 (113) | 10.81 | 12.55 | 6.62 | 6.52 | 28.09 | –1.08 | .288 | .384 |
| Muscle atrophy | 14 (113) | 10.77 | 13.57 | 6.62 | 6.25 | 17.39 | –1.56 | .137 | .274 |
| Fasciculations | 26 (113) | 10.26 | 13.96 | 6.53 | 6.17 | 43.11 | –2.64 | .011* | .046* |
| Myoclonus | 20 (113) | 11.14 | 11.00 | 6.64 | 6.65 | 27.75 | 0.09 | .933 | .933 |
| Rigidity | 2 (113) | 10.96 | 19.50 | 6.55 | 4.95 | 1.06 | –2.40 | .239 | .384 |
| Chorea | 3 (113) | 10.86 | 20.33 | 6.50 | 3.79 | 2.33 | –4.17 | .040* | .092 |
| Dystonia | 16 (113) | 10.91 | 12.38 | 6.60 | 6.72 | 20.08 | –0.81 | .427 | .488 |
| Resting tremor | 2 (113) | 11.30 | 1.00 | 6.52 | 1.41 | 1.91 | 8.75 | .015* | .047* |
| Sensory symptoms | 89 (113) | 9.71 | 11.49 | 7.01 | 6.49 | 34.36 | –1.12 | .268 | .384 |
| Brainstem oculomotor signs | 57 (113) | 9.54 | 12.67 | 7.14 | 5.69 | 104.92 | –2.58 | .011* | .046* |
| Urinary dysfunction | 48 (113) | 9.74 | 12.98 | 6.93 | 5.70 | 109.67 | –2.72 | .008** | .046* |
| Cognitive impairment | 17 (113) | 10.86 | 12.53 | 6.81 | 5.28 | 26.43 | –1.14 | .264 | .384 |

Symbol “+” indicates presence of the respective non-ataxia sign. Symbol “–” indicates absence of the respective non-ataxia sign. \**P*<sub>FDR</sub> < .05. \*\**P*<sub>FDR</sub> < .01. \*\*\**P*<sub>FDR</sub> < .001.

## Supplementary figures

**Supplementary Figure 1.**
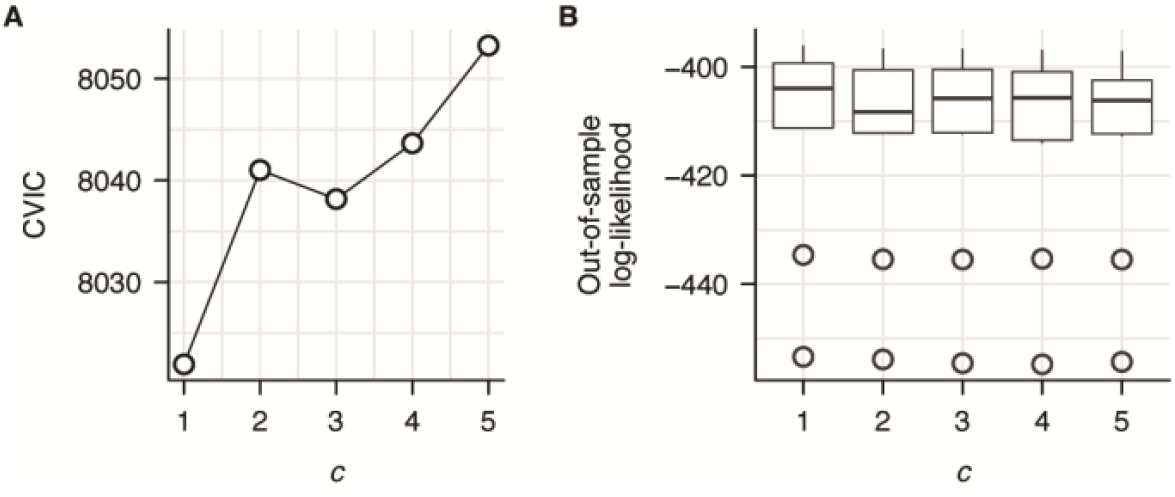
Model selection criteria from ten-fold cross-validation. Both (**A**) cross-validation information criterion (CVIC) and (**B**) out-of-sample log-likelihood clearly indicated that a model for *c* = 1 atrophy subtypes in SCA3 explains the provided data most efficiently.

**Supplementary Figure 2.**
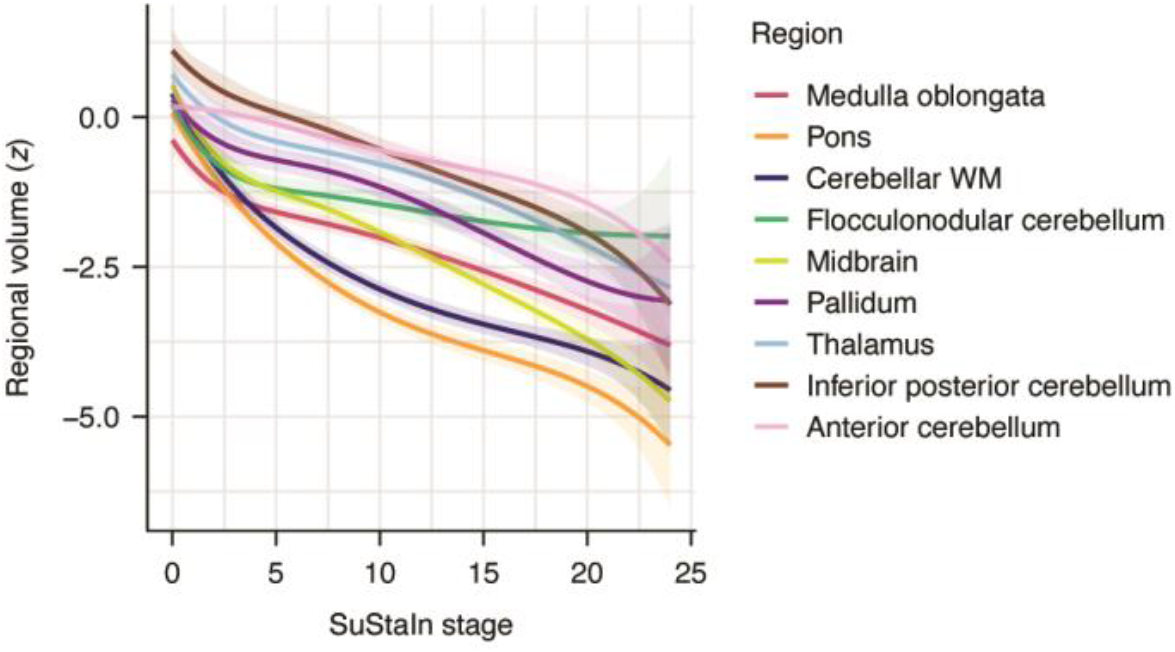
Monotone penalized cubic regression splines of *z*-scored regional volumes across SuStaIn stages. One knot was placed SuStaIn stage = 8, which was the median SuStaIn stage in the present sample. The predicted values of regional volumes displayed on the y-axis are visualized in **Figure 1**.

**Supplementary Figure 3.**
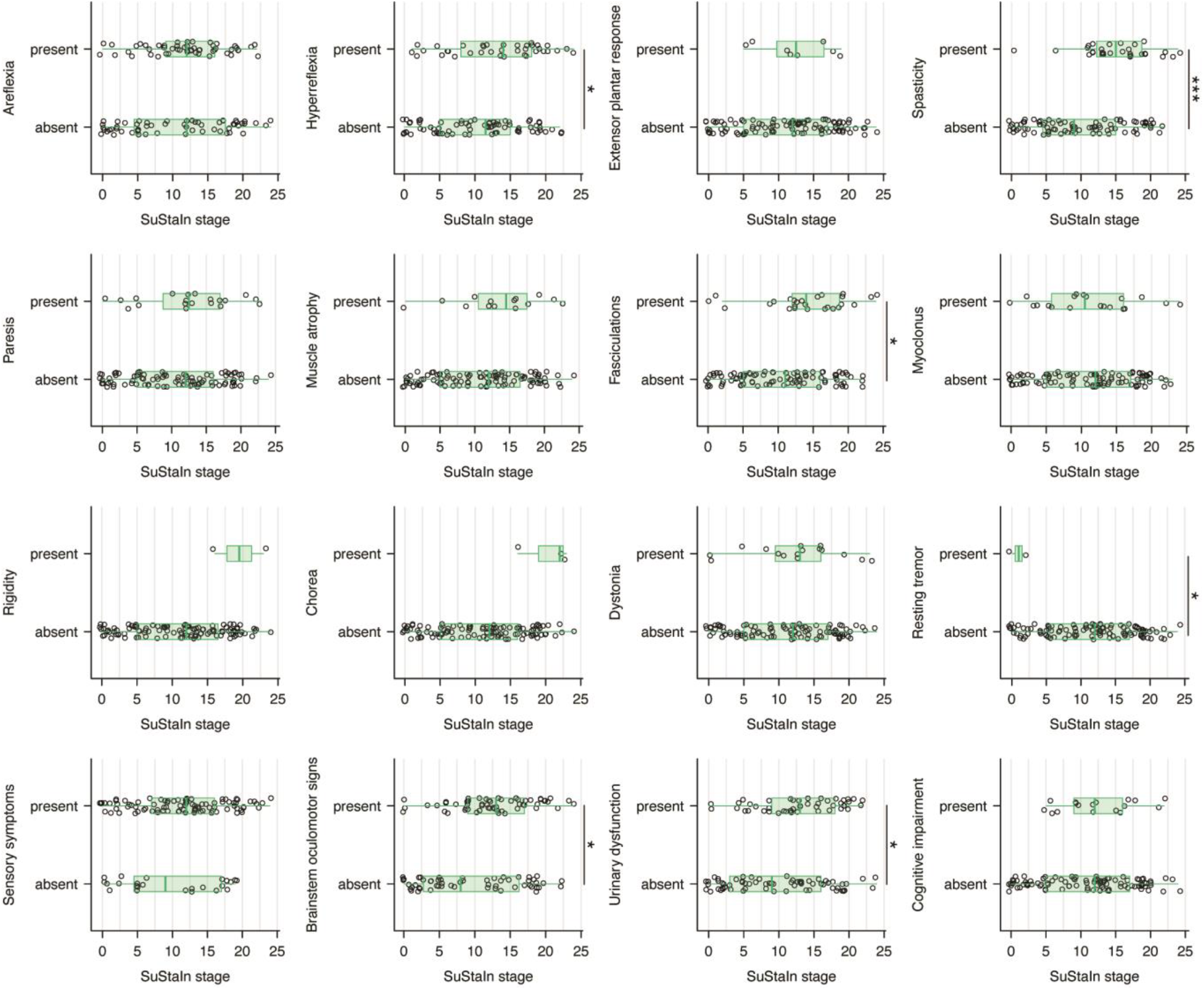
Distributions of SuStaIn stage in SCA3 mutation carriers stratified by the presence or absence of different non-ataxia signs. \**P*_FDR_ < .05. ** *P*_FDR_ < .01. *** *P*_FDR_ < .001.

## Notes

### Author Declarations

Ethics committee/IRB of the University Hospital Bonn gave ethical approval for this work. Ethics committee/IRB of the University of Campinas gave ethical approval for this work. In addition, the ethics committee/IRB of the Xiangya Hospital, Central South University in China (Reference number: 202310206) gave ethical approval for this work. Ethics committee/IRB of the Radboud University Medical Center Nijmegen gave ethical approval for this work. Ethics committee/IRB of the Salpetrier University Hospital Paris gave ethical approval for this work. Ethics committee/IRB of the University Hospital Tuebingen gave ethical approval for this work. Ethics committee/IRB of the UCL London gave ethical approval for this work.Ethics committee/IRB of the University of Minnesota gave ethical approval for this work. Ehtics committee/IRB of the University Hospital Essen gave ethical approval for this work. Ethics committee/IRB of the University Hospital Heidelberg gave ethical approval for this work. Ethics committee/IRB of the University Medical Center Groningen gave ethical approval for this work. Ethics committee/IRB of the Johns Hopkins University School of Medicine, Baltimore, gave ethical approval for this work, as well as the Massachusetts General Hospital. Ethics committee/IRB of the RWTH Aachen gave ethical approval for this work. Ethics committee/IRB of the University of Cantabria gave ethical approval for this work.

### Summary of Updates

Revised version with the correction of a typo of in the Surname of the first author.

